# Combined Infection Control Interventions Protect the Essential Workforce from Occupationally-Acquired SARS-CoV-2 during Produce Production, Harvesting and Processing Activities

**DOI:** 10.1101/2022.04.06.22273125

**Authors:** D. Kane Cooper, Julia S. Sobolik, Jovana Kovacevic, Channah M. Rock, Elizabeth T. Sajewski, Jodie L. Guest, Ben A. Lopman, Lee-Ann Jaykus, Juan S. Leon

**Affiliations:** Rollins School of Public Health, Emory University, Atlanta, GA, 30322, USA; Food Innovation Center, Oregon State University, Portland, OR, 97209, USA; Department of Soil, Water and Environmental Science, University of Arizona, Tucson, AZ 85721, USA; Food, Bioprocessing and Nutrition Sciences, North Carolina State University, Raleigh, NC, 27695, USA

**Keywords:** COVID-19, Quantitative Microbial Risk Assessment (QMRA), Vaccinations, Non-Pharmaceutical Interventions (NPIs), Produce Industry Worker

## Abstract

Essential food workers experience an elevated risk of SARS-CoV-2 infection due to prolonged occupational exposures (e.g., frequent close contact, enclosed spaces) in food production and processing areas, shared transportation (car or bus), and employer-provided shared housing. The purpose of this study was to evaluate the impact of combined food industry interventions and vaccination on reducing the daily cumulative risk of SARS-CoV-2 infection for produce workers. Six linked quantitative microbial risk assessment models were developed in R to simulate daily scenarios experienced by a worker. Standard industry interventions (2 m physical distancing, handwashing, surface disinfection, universal masking, increased ventilation) and two-dose mRNA vaccinations (86–99% efficacy) were modeled individually and jointly to assess risk reductions. The infection risk for an indoor (0.802, 95% Uncertainty Interval [UI]: 0.472–0.984) and outdoor (0.483, 95% UI: 0.255–0.821) worker was reduced to 0.018 (93% reduction) and 0.060 (87.5% reduction) after implementation of combined industry interventions. Upon integration of these interventions with vaccination, the infection risk for indoor (0.001, 95% UI: 0.0001–0.005) and outdoor (0.004, 95% UI: 0.001–0.016) workers was reduced by ≥99.1%. Food workers face considerable risk of occupationally-acquired SARS-CoV-2 infection without interventions; however, consistent implementation of key infection control measures paired with vaccination effectively mitigates these risks.

**Synopsis:** Bundled interventions, particularly if they include vaccination, produce significant reductions (>99%) in SARS-CoV-2 infection risk for essential food workers.

**Graphical Abstract:** 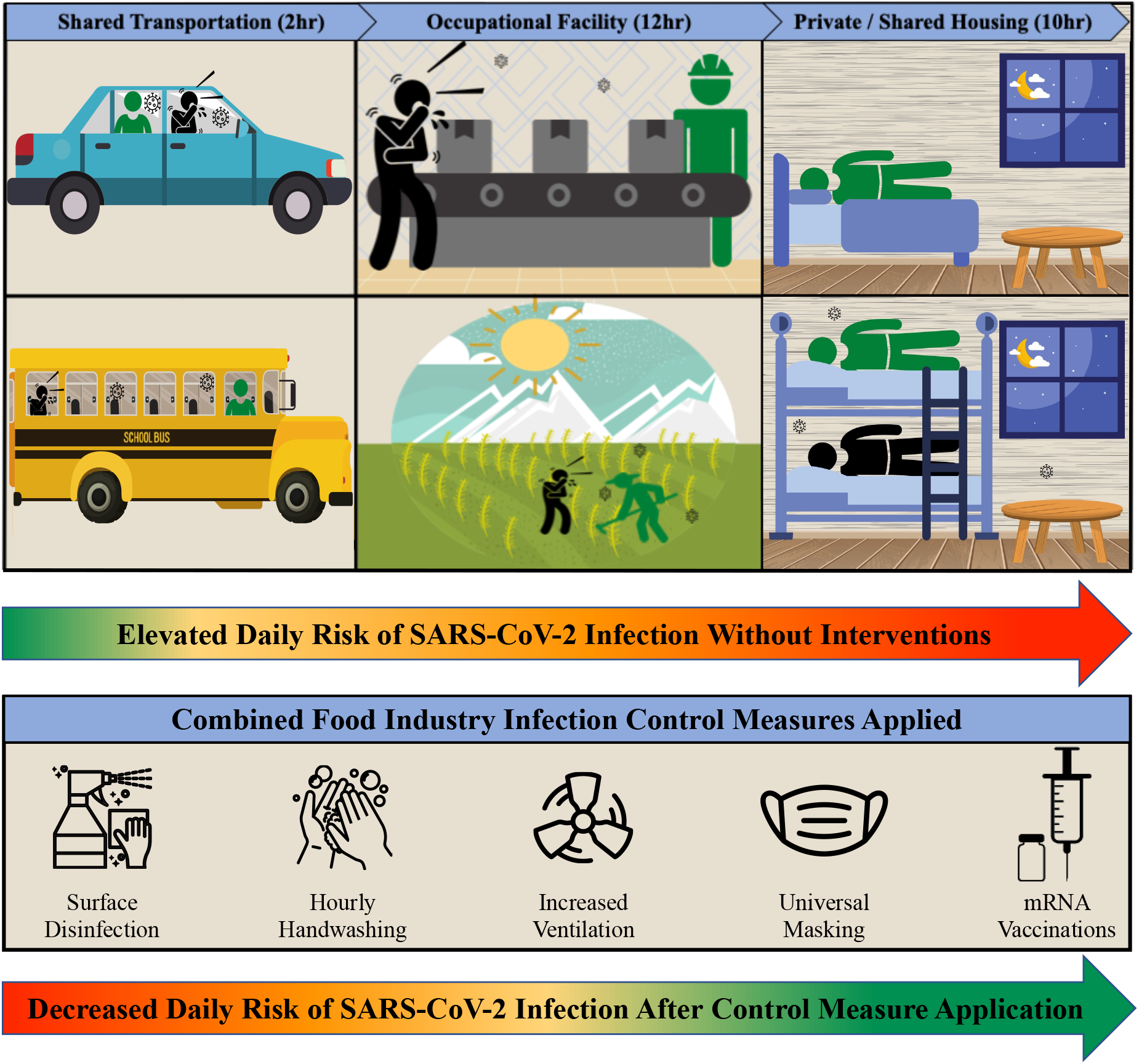

## 1. Introduction

Essential food worker populations have been disproportionately impacted by severe acute respiratory syndrome coronavirus 2 (SARS-CoV-2) illness and death^1–5^. For instance, Canadian migrant farmworkers experienced a 20-fold higher incidence of SARS-CoV-2 infections relative to the general population (July, 2020)^6^. Similarly, food and agricultural workers in California, USA experienced a 39% increase in excess mortality compared to a 12% increase for workers in non-essential sectors (March – November, 2020)^7^. Workplace practices such as close proximity and long shifts may be contributing factors to this increased infection risk^1,8^. Additional exposures may also occur among food workers during shared transportation (carpooling, work bus)^9–11^ and in employer-provided housing^12,13^ (e.g., crowding ^14–17^, poor ventilation^16–18^). Protecting essential food workers from SARS-CoV-2 is necessary to reduce the burden of disease among this often understudied^19^ and marginalized population, and to ensure stability in the global food supply chain^20,21^.

Global agricultural organizations (Food and Agriculture Organization of the United Nations^22,23^, U.S. Department of Agriculture^24^), and food trade associations (Global Cold Chain Alliance^25^, American Frozen Food Institute^26^) have recommended integrating SARS-CoV-2 infection control strategies (e.g., distancing, masking, symptom screening, vaccination, ventilation) to protect food workers. Despite the demonstrated effectiveness of these interventions^27–36^, their impact in protecting food worker populations has been poorly characterized^37^. Implementation of physical barriers and universal masking in 13 Nebraskan meat processing facilities resulted in a significant reduction (62%) in the incidence of COVID-19 among workers^28^. In an indoor food manufacturing scenario, stochastic quantitative microbial risk assessment (QMRA) modeling^38^ revealed that after an 8h work exposure, close contact (1 m) transmission yielded the greatest infection risk (0.98) as compared to worker distances of 2 m (0.15) and 3 m (0.09). Our study found that strategies mitigating respiratory particle exposure (e.g., masking, distancing), paired with worker vaccination reduced infection risk below 1.0%. Similar strategies are also effective at protecting food workers from fomite-mediated transmission during cold-chain packaging and transport activities^39^. While these studies advance the evidence-base of effective risk mitigation strategies in specific food worker scenarios (indoor manufacturing, cold-chain etc.), the unique conditions experienced by much of this workforce (e.g., shared transportation, employer-provided housing) means that daily infection risks may be further impacted by circumstances outside of the production fields or processing plants.

The purpose of this study was to evaluate the impact of combined food industry interventions and vaccination on reducing the daily cumulative risk of SARS-CoV-2 infection among essential produce production and processing workers. This work highlights the elevated risk of infection experienced by this segment of the essential workforce and can inform best practices by the produce industry to maintain the health and wellbeing of their workers.

## 2. Materials and Methods

### 2.1 Model Overview

The daily cumulative infection risk was modeled for a susceptible produce worker exposed to a symptomatic SARS-CoV-2 infected co-worker throughout all representative daily work scenarios (Figure 1). Susceptibility was defined herein as an individual with no underlying health conditions or prior infections altering their level of susceptibility. These daily work scenarios were identified through discussions with industry trade associations, including the American Frozen Food Institute (AFFI) and the Produce Marketing Association (PMA); research and extension experience provided by representatives of the Arizona Cooperative Extension Organization, the Western Regional Center to Enhance Food Safety, and the Emory Farmworker project; and our past research on farms and packing facilities along the United States and Mexico border^40–42^. Each scenario was modeled as an independent QMRA and expanded from previous modeling work of SARS-CoV-2 transmission in an indoor food manufacturing scenario^38^. The SARS-CoV-2 exposure dose was combined across each work scenario experienced by the susceptible produce worker to estimate the daily infection risk.

**Figure 1.**
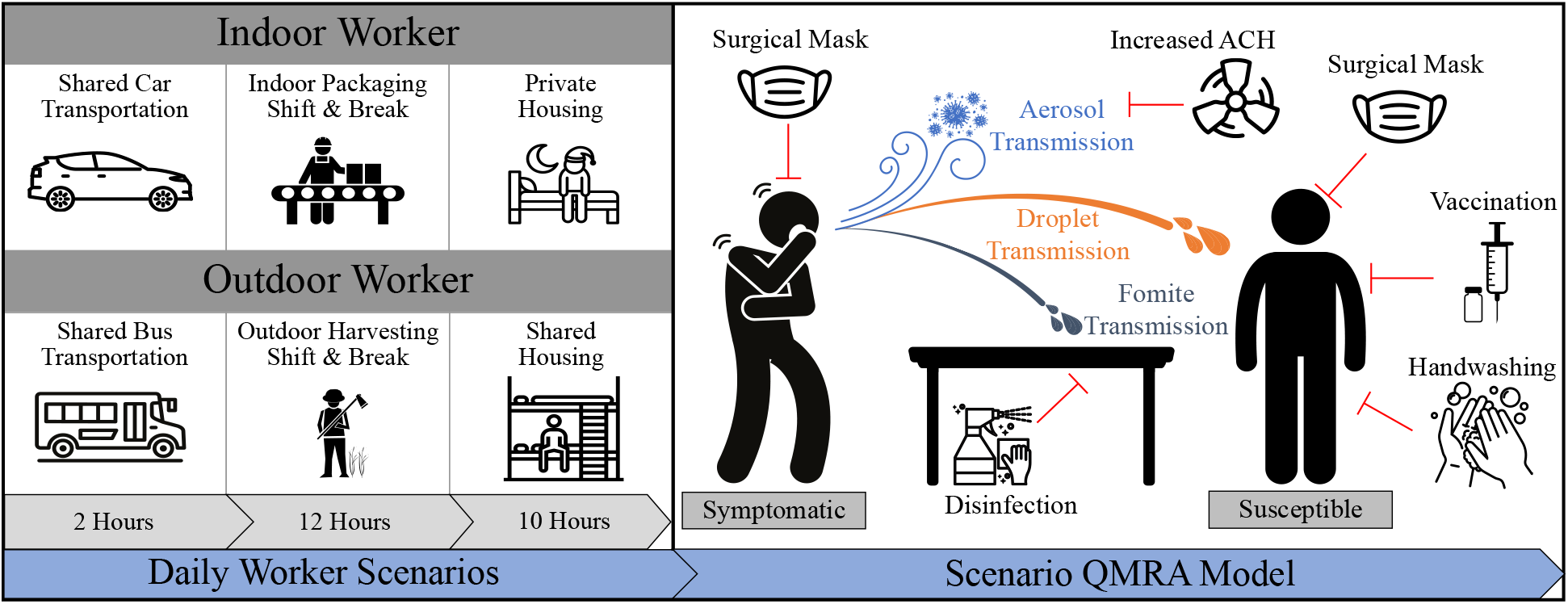
Scenarios impacting daily cumulative risks of SARS-CoV-2 exposure for an indoor and outdoor produce worker. *Left – Daily Worker Scenarios*. Simulations were conducted in which workers were engaged in three overarching daily scenarios: shared transportation, working shift, and shared housing (see Methods). Simulations were done in which the workers take a 1h break during their work shift in a break room (indoor) or inside their bus transportation (outdoor). ***Right – Scenario QMRA Model***. Each scenario simulates the risk of SARS-CoV-2 transmission (aerosol-, droplet-, and fomite-mediated), from an infected, coughing co-worker, and the impact of infection control strategies (red lines), adjusted for the context of each scenario (see Methods).

International (World Health Organization) and domestic (U.S. Centers for Disease Control and Prevention, U.S. Food and Drug Administration, U.S. Occupational Safety and Health Administration) guidance for controlling SARS-CoV-2 in the food industry, along with insight from authors (J.K., C.M.R.) and experts at AFFI, was used to inform the interventions applied across each modeled scenario. Interventions included the individual and combined impact of physical distancing, handwashing and surface disinfection, universal masking, increased ventilation, and SARS-CoV-2 vaccination on reducing infection risk for a susceptible worker. The model outcomes include: 1) the scenario-specific, as well as the daily cumulative SARS-CoV-2 infection risk for a susceptible produce worker quantified across three viral transmission pathways (aerosol, droplet, and fomite-mediated); and 2) the individual and combined impact of recommended infection control strategies on reducing the daily cumulative risk of SARS-CoV-2 infection for a susceptible produce worker.

### 2.2 Sequential work scenarios for an indoor and outdoor produce workers

Daily scenarios experienced by produce industry workers were categorized into “indoor” and “outdoor” work scenarios (Figure 1) based on the activity-specific differences and risk profiles of each worker population and industry expert input. For example, while both indoor and outdoor workers engage in shared transportation to and from work^19,43–45^, the specific mode of transportation (i.e., shared car [indoor] or bus [outdoor]) was determined through conversations with produce industry managers and farmworker extension specialists. For an indoor susceptible worker, we assumed that 2 h would be spent in shared car transportation, followed by a 12 h shift in an indoor produce packaging facility (11 h working, 1 h break), and 10 h in private housing. The indoor worker was assumed to be exposed to an infected worker in all scenarios except for in the private housing. For an outdoor susceptible worker, we assumed that 2 h would be spent in shared bus transportation, followed by a 12 h shift in an outdoor produce harvesting field (11 h working, 1 h break), and 10 h in shared housing. Exposure to an infected worker, for the outdoor worker, was assumed in each of the work scenarios. Details regarding the parameterization of these scenarios can be found in the **Supporting Information**.

### 2.3 QMRA model framework for each scenario

Expanding upon our previous SARS-CoV-2 QMRA model, we quantified the risk of infection across three transmission pathways (aerosols, droplets, fomites) for both indoor and outdoor food workers^38^. Each modeled scenario assumed a single infected worker either coughed (symptomatic) or breathed (asymptomatic) virus-laden respiratory aerosols (particle diameter <50 μm) and droplets (particle diameter 50–750 μm) that could infect a susceptible worker. Aerosols were assumed to travel distances beyond 3 m from their source and were homogenously distributed in the environment^46–48^, while droplets were assumed to travel <3 m from their source and fell to adjacent surfaces based on particle size and simulated distance traveled^49–51^. Fomite contamination occurred through infectious aerosols and droplets falling onto surfaces based on their settling velocities and gravitational trajectories^52^. For each modeled scenario, we incorporated specific dimensions, ventilation rates, fomite surface materials, and viral decay rates as described below.

### 2.4 SARS-CoV-2 Risk Characterization

The models were constructed in R (version 4.0.3; R Development Core Team; Vienna, Austria) with 10,000 iterations, using literature-derived specified parameters and probability distributions. Each model parameter used to inform the SARS-CoV-2 viral transmission pathways was grouped into three categories and summarized in **Supporting Table 1A-C**. These classifications include: 1) viral shedding through coughing respiratory events; 2) fomite-mediated transmission and dose-response parameters; and 3) risk mitigation interventions (infection control interventions and vaccinations). Scenario-specific parameters, such as the volume of space modeled, temperature and relative humidity, baseline air exchange rate (ACH), and fomite-specific viral decay parameters are in **Supporting Table 2**.

The scenario-specific risk of SARS-CoV-2 infection to a susceptible produce worker was calculated by summing the viral dose across each transmission pathway (aerosol, droplet, fomite) for each scenario (shared transportation, work shift, work break, shared housing). The daily cumulative risk of infection to a susceptible produce worker was calculated by summing the dose across scenarios specific to either an indoor or outdoor produce worker. Both the scenario-specific and daily cumulative viral doses were then converted into risk estimates using the SARS-CoV-1 dose-response parameter, as described below.

Aerosol and droplet exposures to the susceptible worker were calculated from the concentration of virus (*C*_*t*_) at time *t*, the deposition fraction of particles into the lung mucosa (*L*_*dep*_), the inhalation rate (*I*_*R*_), and the duration of exposure (*E*_*t*_) as follows:

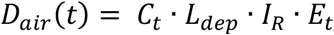

The fomite-mediated viral exposure was based on the frequency of hand-to-face contacts (*H*_*face*_), the ratio of finger (*F*_*sa*_)-to-hand (*H*_*sa*_) surface area, the concentration of virus on the hand at each time *t* (*C*_*hand*_), the fraction of pathogens transferred from the hand to the facial mucosal membrane (*F*_*23*_), and the exposure duration in hours as follows:

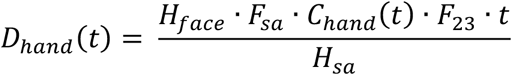

These viral doses were then combined to generate scenario-specific SARS-CoV-2 exposures. For instance, the viral dose for the 11 h indoor processing facility shift was calculated by:

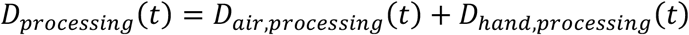

The daily cumulative viral dose for an indoor (*D*_*indoo*r_) or outdoor (*D*_*outdoor*_) produce worker was calculated by combining the scenario-specific viral doses (see **Supporting Information**) relevant to each worker as follows:

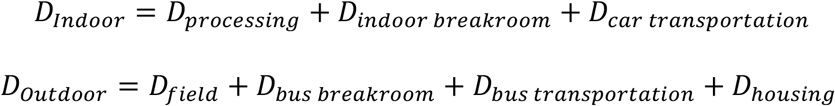

The probability of SARS-CoV-2 infection to a susceptible produce worker from an infected worker was calculated using an exponential dose-response model (k_risk_) based on pooled SARS-CoV-1 laboratory data using the intranasal inoculation of mice models^53^. For example, the scenario-specific infection risk associated with the shared car transportation scenario (R_car transportation_) and daily cumulative risk for an indoor produce worker (R_Indoor_) were calculated by:

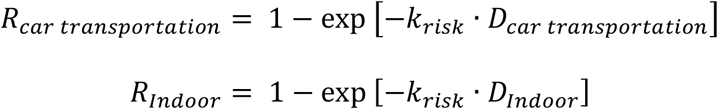

Results are presented as the median risk values with the 95% uncertainty interval (UI), which is comprised of the 2.5 and 97.5 percentiles.

### 2.5 Selection of Infection Control Strategies for SARS-CoV-2

Recommended infection control strategies (e.g., physical distancing, handwashing and surface disinfection, universal mask wearing, increased air exchange rates, and vaccination) were identified from a systematic review of 1,847 United Nations and English-speaking government websites to distill global guidance, guidelines, or recommendations to prevent food workers from acquiring COVID-19^54^. These strategies were then vetted with our trade association partners (AFFI, PMA) and extension co-authors (J.K. and C.M.R.). The impact of interventions (individually or combined) was evaluated based on the reduction in infection risk (%) relative to the baseline scenario with no interventions.

Increasing the distance (physical distancing) between workers from 1 m to 2 m, based on CDC and OSHA guidance^55^, was analyzed in the indoor packing facility and outdoor harvesting field scenarios. The impact of handwashing and surface disinfection on reducing the risk of SARS-CoV-2 infection was analyzed in the indoor packing facility and outdoor harvesting field scenarios. Here, we assumed hourly handwashing (2-log_10_ viral removal)^56^ per handwashing event. Surface disinfection (3-log_10_ viral removal) using EPA N-list of SARS-CoV-2 viral disinfectants^57^ was assumed to occur twice per work shift (4 h, 8 h). The impact of various facemask materials (cloth, surgical) and double masking (surgical mask followed by cloth mask) was assessed across all scenarios except for the indoor and outdoor 1 h breaks to allow for eating, and in the 8 h in the shared residential scenario to allow for sleeping. The range in efficacy for each facemask material can be found in Supporting Table 1C.

Scenario-specific increases in air exchange rate (ACH) based on particulate exposure studies were assessed across each modeled scenario except for the 11 h outdoor harvesting field shift and 1 h bus transportation. For the shared car transportation scenario, the baseline ACH of 2.55 h^-1^ (95% UI: 0.98–4.10) was increased to 29.1 h^-1^ (95% UI: 7.67–50.6), which was representative of a 2005 Ford Taurus mid-size car traveling from 0–50 mph with all windows open^58^. For both the indoor packaging facility and associated breakroom, the ACH was increased from 0.1 h^-1^ to 6.0 h^-1^ based on surveys and conversations with food industry managers (data not shown). For the shared bus transportation scenario, the baseline ACH of 11.1 h^-1^ (95% UI: 8.96–13.4) was increased to 21.6 h^-1^ (95% UI: 10.5–37.3), which was representative of a school bus driving under realistic conditions with all windows open^59^. For the shared residential scenario, the baseline ACH of 0.35 h^-1^ (point estimate) was increased to 3.9 h^-1^ (95% UI: 2.08–5.72), which representative of opening a window to increase natural ventilation in an apartment-style scenario^60^. Additional information on the scenario-specific parameters can be found in **Supporting Table 2**.

Finally, we evaluated the impact of two vaccination scenarios (sub-optimal and optimal vaccine efficacies) on reducing the daily cumulative risk of SARS-CoV-2 infection for a susceptible worker. We assumed that across all modeled scenarios, only the susceptible worker would be vaccinated. For the sub-optimal vaccine efficacy scenario, the mean vaccine efficacy was 72.0% (95% UI: 64.6–79.6%) and represented either receiving one dose of the Johnson & Johnson vaccine, one of the two-dose vaccine series, a reduced vaccine efficacy to novel variants, or incomplete immunity due to a prior SARS-CoV-2 infection^29,31,32^. For the full immunity scenario, the mean vaccine efficacy was 92.3% (95% UI: 86.3–98.7%) and represented two doses (≥14 days) of Pfizer-BioNTech or Moderna-NIAD mRNA series vaccines^33,34^ by the susceptible worker. The impact of booster vaccinations was not assessed; however, receiving three doses of the mRNA vaccines has been shown to provide a mean vaccine efficacy of 94.0%, aligning with the range represented in the optimal vaccine efficacy scenario^61^.

### 2.6 Sensitivity Analysis

Sensitivity analyses were conducted for each modeled scenario to determine the most influential parameters in estimating the SARS-CoV-2 viral exposure dose. Parameters identified as being most influential in the final cumulative dose estimate were reported as Spearman rank correlational coefficients using the “tornado” function in the mc2d R package^62^. To investigate the propagation of variability and uncertainty throughout the models, the “mcratio” function was used to calculate the variability and overall uncertainty ratio for each modeled parameter. More information on the modeled parameters, their distributions, and assessing model stability can be found in the **Supporting Information**.

### 2.7 Data Availability

The code developed and utilized throughout this analysis will be available through GitHub upon acceptance.

## 3. Results

### 3.1 Scenario-specific and cumulative daily SARS-CoV-2 infection risks for an indoor produce worker with and without infection control measures

For an indoor produce worker, we investigated the individual and cumulative SARS-COV-2 infection risk to a susceptible worker across four daily scenarios: shared car transportation (2 h), an indoor facility work shift (11 h), break in the indoor facility breakroom (1 h), and private housing (10 h, Figure 1). With no infection control strategies applied, the lowest risk scenario was the time spent in the indoor breakroom (Risk: 0.021, 95% Uncertainty Interval [UI]: 0.008– 0.053) (Table 1), whereas the highest risk scenario was during the 11 h enclosed work shift at 1 m distancing (1.00, 95% UI: 0.977–1.00). Within the 11 h enclosed work shift scenario when distancing increased to 2 m, the risk decreased by 61.3% (0.387, 95% UI: 0.162–0.731).

**Table 1.** Cumulative Risk and Risk Reduction (%) of SARS-CoV-2 Infection for a Susceptible Indoor Produce Worker Exposed to an Infected Co-Worker

| Infection Control Strategy | Shared Car Transportation | Work Shift Distancing |  | Break in Breakroom | Private Housing | Daily Cumulative Risk* Distancing |  |
| --- | --- | --- | --- | --- | --- | --- | --- |
|  |  | 1m | 2m |  |  | 1m | 2m |
| None<sup>†</sup> | 0.664<br>(0.325–0.943) | 1.00<br>(0.977–1.00) | 0.387<br>(0.162–0.731) | 0.021<br>(0.008–0.053) | None | 1.00<br>(0.992–1.00) | 0.802<br>(0.472–0.984) |
| Handwashing & Surface Disinfection<sup>‡</sup> | No | 1.00<br>(0.00%) | 0.361<br>(6.59%) | No | None | 1.00<br>(0.00%) | 0.792<br>(1.25%) |
| Cloth Mask<sup>‡</sup> | 0.148<br>(77.7%) | 0.845<br>(15.5%) | 0.080<br>(79.4%) | No | None | 0.876<br>(12.4%) | 0.226<br>(71.8%) |
| Surgical Mask<sup>‡</sup> | 0.099<br>(85.1%) | 0.694<br>(30.6%) | 0.056<br>(85.5%) | No | None | 0.735<br>(26.5%) | 0.158<br>(80.3%) |
| Double Mask<sup>‡</sup> | 0.023<br>(96.5%) | 0.274<br>(72.6%) | 0.018<br>(95.4%) | No | None | 0.320<br>(68.0%) | 0.048<br>(94.0%) |
| Increased Air Exchange<sup>‡</sup> | 0.125<br>(81.2%) | 0.896<br>(10.4%) | 0.049<br>(87.2%) | 0.004<br>(81.0%) | None | 0.916<br>(8.40%) | 0.181<br>(77.4%) |

<sup>†</sup> Values represent Median (95% Uncertainty Interval) SARS-CoV-2 infection risk.
<sup>‡</sup> Values represent Median (% Reduction) SARS-CoV-2 infection risk compared to no Infection Control Strategy. “No” indicates the Infection Control Strategy was not applied (see Methods). “None” means that there was no risk of infection in the private housing scenario.
\* Assuming there is contact between an infected and susceptible individual in every worker scenario.

However, the 24 h daily cumulative risk remained elevated for both 1 m (1.00, 95% UI: 0.992– 1.00) and 2 m (0.802, 95% UI: 0.472–0.984) distancing. After implementing individual infection control strategies across each modeled scenario, for the 2 m 24 h daily cumulative risk, universal double masking resulted in the greatest reduction to 94.0% (0.048, 95% UI: 0.006–0.316), while surgical masking reduced this risk by 80.3% (0.158, 95% UI: 0.040–0.424), and cloth masking by 71.8% (0.226, 95% UI: 0.075–0.557). For the 2 m 24 h daily cumulative risk, increasing ventilation rates across each modeled scenario resulted in a 77.4% reduction (0.181, 95% UI: 0.078–0.571), while hourly handwashing and surface disinfection twice during the 11 h work shift provided a nominal reduction of 1.25% (0.792, 95% UI: 0.456–0.983). The 24 h daily cumulative risk at 2 m was lower for each intervention assessed when compared to the daily cumulative risk at 1 m.

### 3.2 Scenario-specific and cumulative daily SARS-CoV-2 infection risks for an outdoor produce worker with and without infection control measures

Next, we investigated the individual and cumulative risk to a susceptible outdoor food worker exposed to an infected worker across four daily scenarios: shared bus transportation (2 h), an outdoor field work shift (11 h), a rest break in the bus (1 h), and shared housing (10 h, Figure 1) (Table 2). Without infection control measures, the lowest risk scenario was the time spent in the shared bus transportation (0.010, 95% UI: 0.003–0.031) (Table 2), whereas the highest risk scenario was during the 11 h outdoor field work shift at 1 m distancing (0.316, 95% UI: 0.098– 0.768). Within the 11 h outdoor work shift scenario when distancing increased to 2 m, the risk decreased by 97.5% (0.008, 95% UI: 0.003–0.019). However, the 24 h daily cumulative risk remained elevated for both 1 m (0.483, 95% UI: 0.255–0.821) and 2 m (0.215, 95% UI: 0.095– 0.441). After implementing individual infection control strategies across each modeled scenario, for the 1 m 24 h daily cumulative risk, universal double masking resulted in the greatest reduction to 91.8% (0.039, 95% UI: 0.016–0.140), while surgical masking reduced this risk by 83.4% (0.079, 95% UI: 0.031–0.215), and cloth masking by 76.7% (0.112, 95% UI: 0.048– 0.287). For the 1 m 24 h daily cumulative risk, increasing ventilation rates in the shared bus transportation and shared housing scenarios resulted in a 16.2% reduction (0.404, 95% UI: 0.181–0.802) in the 24 h daily cumulative risk at 1 m, while hourly handwashing and surface disinfection twice during the 11 h outdoor work shift provided a nominal reduction in the 24 h cumulative risk at 1 m of 0.15% (0.482, 95% UI: 0.254–0.820). The 24 h daily cumulative risk at 2 m was lower for each intervention assessed when compared to the daily cumulative risk at 1 m.

**Table 2.** Cumulative Risk and Risk Reduction (%) of SARS-CoV-2 Infection for a Susceptible Outdoor Produce Worker Exposed to an Infected Co-Worker

| Infection Control Strategy | Shared Bus Transportation | Work Shift Distancing |  | Rest Break in the Bus | Shared Housing | Daily Cumulative Risk* Distancing |  |
| --- | --- | --- | --- | --- | --- | --- | --- |
|  |  | 1m | 2m |  |  | 1m | 2m |
| None<sup>†</sup> | 0.010<br>(0.003–0.031) | 0.316<br>(0.098–0.768) | 0.008<br>(0.003–0.019) | 0.012<br>(0.005–0.027) | 0.190<br>(0.075–0.411) | 0.483<br>(0.255–0.821) | 0.215<br>(0.095–0.441) |
| Handwashing & Surface Disinfection<sup>‡</sup> | No | 0.315<br>(0.21%) | 0.007<br>(4.47%) | No | No | 0.482<br>(0.15%) | 0.2148<br>(0.11%) |
| Cloth Mask<sup>‡</sup> | 0.001<br>(85.5%) | 0.055<br>(82.7%) | 0.001<br>(83.4%) | No | 0.036<br>(80.9%) | 0.112<br>(76.7%) | 0.052<br>(76.0%) |
| Surgical Mask<sup>‡</sup> | 9.3x10<sup>-4</sup><br>(90.7%) | 0.034<br>(89.2%) | 8.5x10<sup>-4</sup><br>(88.9%) | No | 0.026<br>(86.5%) | 0.079<br>(83.4%) | 0.040<br>(81.4%) |
| Double Mask<sup>‡</sup> | 2.2x10<sup>-4</sup><br>(97.8%) | 0.009<br>(97.2%) | 2.7x10<sup>-4</sup><br>(96.5%) | No | 0.012<br>(93.7%) | 0.039<br>(91.8%) | 0.026<br>(88.1%) |
| Increased Air Exchange<sup>‡</sup> | 0.005<br>(45.2%) | N/A | N/A | No | 0.098<br>(48.6%) | 0.404<br>(16.2%) | 0.122<br>(43.1%) |

<sup>†</sup> Values represent Median (95% Uncertainty Interval) SARS-CoV-2 infection risk.
<sup>‡</sup> Values represent Median (% Reduction) SARS-CoV-2 infection risk compared to no Infection Control Strategy. “No” indicates the Infection Control Strategy was not applied (see Methods). “N/A” means that it is not applicable to apply increased air exchange in an outdoor environment.
\* Assuming there is contact between an infected and susceptible individual in every worker scenario.

### 3.3 Impact of combined infection control strategies and vaccination on the daily cumulative risk of SARS-CoV-2 infection for an indoor produce worker

We then investigated the impact of combined infection control measures with and without vaccination on the daily cumulative SARS-CoV-2 risk for the indoor produce worker (Figure 2). Without vaccination, combining hand hygiene, surface disinfection, universal surgical mask usage, and increased ventilation strategies across the indoor work scenarios reduced the daily infection risk by 93.0% (0.018, 95% UI: 0.004–0.098). These combined infection control measures were found to be more effective than vaccination alone (sub-optimal or optimal vaccine efficacies). In the absence of infection control measures, daily infection risk was reduced by 73.2% (sub-optimal vaccine efficacy risk: 0.215, 95% UI: 0.120–0.324) and 92.7% (optimal vaccine efficacy risk: 0.059, 95% UI: 0.007–0.132). Combining infection control strategies with vaccination further reduced the daily infection risks by 99.4% (sub-optimal vaccine efficacy: 0.005, 95% UI: 0.001–0.033), and 99.8% (optimal vaccine efficacy: 0.001, 95% UI: 0.0001–

**Figure 2.**
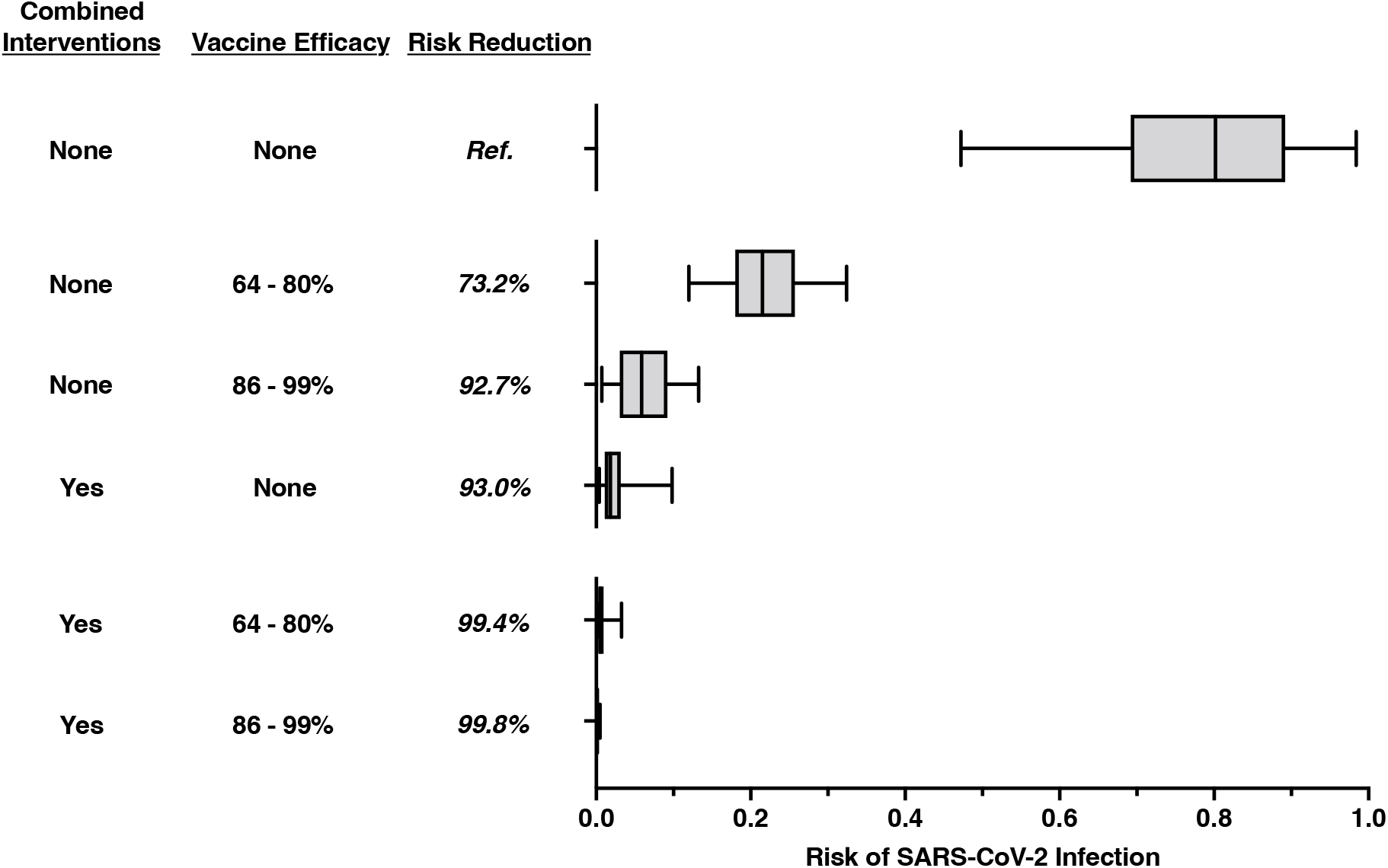
Indoor produce workers experience the greatest reduction in SARS-CoV-2 infection risk when vaccinations are applied in combination with recommended infection control strategies. Combined infection control interventions represent simultaneous surgical mask usage, hourly handwashing, surface disinfection (twice per work shift), and an increased air exchange rate per scenario. For the sub-optimal vaccine efficacy intervention, the susceptible worker was assumed to have a 64–80% reduction (mean: 72.0%, 95% UI: 64.6–79.6%) in infection risk, representative of either receiving one dose of the Johnson & Johnson vaccine, a single dose of the two-dose mRNA vaccine series, or reduced vaccine efficacy against novel SARS-CoV-2 variants^29,31,32^. For the optimal vaccine efficacy intervention, the susceptible worker was assumed to have an 86–99% reduction (mean: 92.3%, 95% UI: 86.3–98.7%) in infection risk which is representative of receiving both doses in the two-dose mRNA vaccine series and is derived from Pfizer-BioNTech and Moderna vaccine effectiveness data^33,34^. Risk reductions were calculated by comparing the risk estimate per intervention scenario against the baseline (no intervention) risk estimate. The top row represents the baseline (reference) daily cumulative infection risk for an indoor produce worker, across the 2 h shared car transportation and 12 h indoor food manufacturing work shift (at 2 m distance) scenario with no infection control strategies applied. The median risk of infection is denoted by the line within each box plot, with error bars representing the 95% uncertainty interval.

0.005).

### 3.4 Impact of combined infection control strategies and vaccination on the daily cumulative risk of SARS-CoV-2 infection for an outdoor produce worker

Similarly, we evaluated the impact of combined infection control measures with and without vaccination on the daily cumulative SARS-CoV-2 infection risk for an outdoor produce worker (Figure 3). Without vaccination, combining hand hygiene, surface disinfection, universal surgical mask usage, and increased ventilation strategies during transportation and in shared housing across the outdoor work scenarios reduced the daily infection risk by 87.5% (0.060, 95% UI: 0.022–0.205). These combined infection control measures were found to be more effective than vaccination with a sub-optimal efficacy, but less effective than vaccination with optimal efficacy. In the absence of infection control measures, daily infection risk was reduced by 72.5% (sub-optimal vaccine efficacy risk: 0.133, 95% UI: 0.065–0.249) and 93.0% (optimal vaccine efficacy risk: 0.034, 95% UI: 0.006–0.087). Combining infection control strategies with vaccination further reduced the daily infection risks by 96.5% (sub-optimal vaccine efficacy: 0.017, 95% UI: 0.006–0.059) and 99.1% (optimal-vaccine efficacy: 0.004, 95% UI: 0.001–0.016).

**Figure 3.**
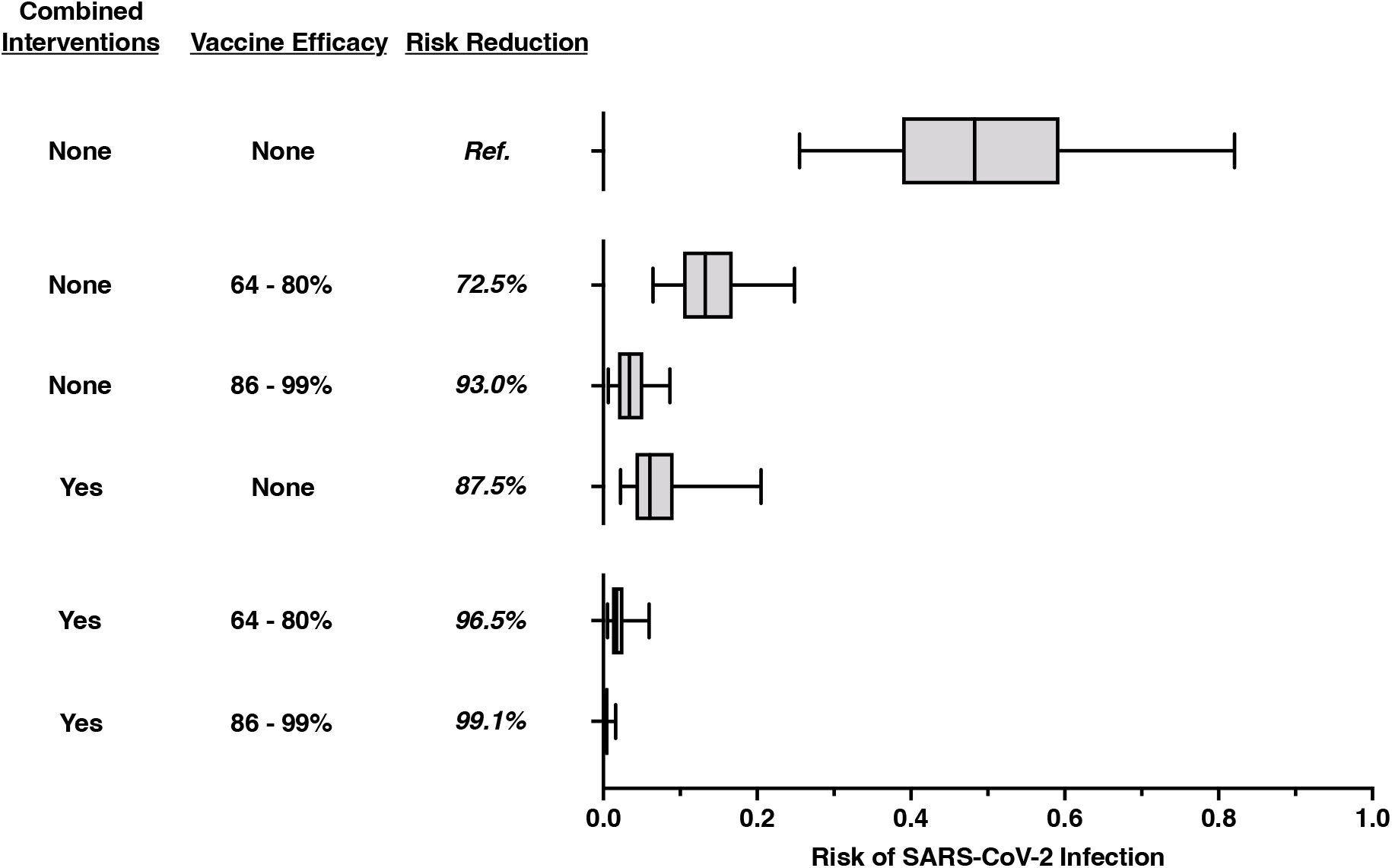
Outdoor produce workers experience the greatest reduction in SARS-CoV-2 infection risk when vaccinations are applied in combination with non-pharmaceutical interventions. Combined infection control interventions represent simultaneous surgical mask usage, hourly handwashing, surface disinfection (twice per work shift), and an increased air exchange rate per scenario. For the sub-optimal vaccine efficacy intervention, the susceptible worker was assumed to have a 64–80% reduction (mean: 72.0%, 95% UI: 64.6–79.6%) in infection risk, representative of either receiving one dose of the Johnson & Johnson vaccine, a single dose of the two-dose mRNA vaccine series, or reduced vaccine efficacy against novel SARS-CoV-2 variants^29,31,32^. For the optimal vaccine efficacy intervention, the susceptible worker was assumed to have an 86–99% reduction (mean: 92.3%, 95% UI: 86.3–98.7%) in infection risk which is representative of receiving both doses in the two-dose mRNA vaccine series and is derived from Pfizer-BioNTech and Moderna vaccine effectiveness data^33,34^. Risk reductions were calculated by comparing the risk estimate per intervention scenario against the baseline (no intervention) risk estimate. The top row represents the baseline daily cumulative risk of infection for an outdoor produce worker, summed across the 2 h shared bus transportation, 12 h outdoor field harvesting shift (at 1 m distance), and 10 h shared housing scenario with no interventions applied. The median risk of infection is denoted by the line within each box plot, with error bars representing the 95% uncertainty interval.

### 3.5 Sensitivity analyses

Spearman rank correlation coefficients were calculated for each scenario to identify the parameters that were the most influential to the final SARS-CoV-2 exposure dose estimate. Across all scenarios, the parameters identified as most influential at increasing virus exposure were the viral shedding rate per hour (ρ_avg_ = 0.95); the infected worker’s salivary virus concentration (ρ_avg_ = 0.81); and the coughing frequency (ρ_avg_ = 0.33). The parameters identified as most influential in decreasing virus exposure across all scenarios were the infected worker’s double masking (ρ_avg_ = -0.73); the susceptible worker’s surgical masking (ρ_avg_ = -0.68); and the susceptible worker’s cloth masking (ρ_avg_ = -0.53) (**Supporting Figure 3A-B**). The shared residential and car transportation scenarios were found to be impacted the most by parameter variability, with overall variability ratios calculated to be 6.06 (aerosol transmission) and 4.60 (droplet transmission), respectively (**Supporting Figure 2A-B**). Finally, the overall uncertainty ratio, representing the combined effect of parameter variability and uncertainty, was largest for the viral dose on a susceptible worker’s hand (4.72) and the Brownian diffusivity (4.34) calculated for aerosol particle exposure (**Supporting Table 3**).

## 4. Discussion

The goal of this study was to estimate the impact of recommended infection control interventions on the daily cumulative SARS-CoV-2 infection risk to produce workers (i.e., working in an indoor facility or open outdoor farm) when exposed to various scenarios (i.e., 1 m or 2 m distancing, shared transportation, with or without shared housing). Among the scenarios assessed for an indoor worker, we found the highest risk to be associated with the 1 m distancing indoor work shift (1.00, 95% UI: 0.977–1.00), followed by shared car transportation (0.664; 95% UI: 0.325–0.943). For an outdoor worker, the highest risk was associated with the 1 m distancing outdoor work shift (0.316, 95% UI: 0.098–0.768), followed by shared housing (0.190, 95% UI: 0.075–0.411). When evaluating the scenario-specific impact of each infection control intervention, we found that double masking provided the greatest protection for the indoor worker, while increased physical distancing between workers provided the greatest protection for outdoor workers. However, the greatest reduction in daily SARS-CoV-2 infection risk was observed when the combined infection control interventions were paired with optimal SARS-CoV-2 vaccinations for both indoor workers (0.001, 99.8% reduction) and outdoor workers (0.004, 99.1% reduction). Despite elevated SARS-CoV-2 exposures throughout daily produce worker activities, this study demonstrates that current risk mitigation strategies when implemented together can effectively protect essential food workers.

Our findings for the indoor packaging facility and outdoor harvesting field are consistent with data from well-documented outbreaks across multiple farms and food processing facilities globally^1,2,21,44,63^, which have highlighted the increased duration of contact and close proximity of workers on assembly lines as factors contributing to an elevated infection risk. In our study, one contributing factor for this elevated infection risk is likely the extended exposure durations (11 h work shift and 1 h break) to a coughing infected worker compared to other scenarios for which exposure durations are in the 1-2 h range. We also explored the impact of physical distancing during the work shifts and found that in the indoor facility and outdoor field, increasing the distance between workers from 1 m to 2 m provided a 61.3% (indoor) and 97.5% (outdoor) infection risk reduction. Consistent with modeling work by Wei et al.,^64^ there is an attenuation in distance traveled by respiratory particles with increasing size: small-diameter (<50 μm) respiratory aerosols can travel beyond 4 m after coughing, whereas 99% of large-diameter droplets (>100 μm) fall to adjacent surfaces within 2 m of their point of origin. As demonstrated here and in our previous modeling work^38^, increasing the distance between workers allows for a shift in the predominant transmission route from close-contact droplets and aerosols to exclusively aerosols (<50 μm; responsible for 1.3% of expelled viral dose), further exemplifying the importance of physical distancing to reduce infection risk.

The elevated infection risk found in the shared car transportation scenario is consistent with Ng et al.^65^, who found that sharing a vehicle with an individual infected with SARS-CoV-2 was associated with a 3.05-fold increase in the odds of infection. This finding is likely due to the relatively low volume of air space (2.6 m^3^) within the car, combined with the elevated persistence of infectious aerosols over multiple hours^66^. For example, after increasing the volume inside of the modeled car by three-fold to 7.8 m^3^ and keeping all other parameters constant, we observed a 53.2% reduction in the 2 h infection risk (0.311, 95% UI: 0.130–0.616) attributed to volume alone. Given our assumption of a well-mixed environment in which the persistent aerosol particles are homogeneously distributed, one would expect aerosol exposures and subsequent infection risks to decrease as the air volume within the scenario increases. Similarly, conditions such as poor ventilation and small and enclosed employer-provided housing have been reported in various outbreaks among outdoor produce workers^14–18^. For example, in July 2020, a California farmworker housing facility with five workers per room documented a 204-person SARS-CoV-2 outbreak^67^. As others have reported^68–71^, the smaller the size of the enclosed space and the more individuals in that space, the greater the risk of SARS-CoV-2 transmission. Taken together, these scenarios highlight the interconnected relationship between duration of exposure, distancing between individuals, and the size/volume of the enclosed space, on SARS-CoV-2 infection risk among indoor produce workers.

Increased ventilation, relative to other infection control strategies, resulted in the greatest reduction in daily infection risk for an indoor worker (77.4% reduction at 2 m), while double masking provided the largest protective effect for both indoor (94.0% reduction at 2 m) and outdoor workers (91.8% reduction at 1 m). Our findings are consistent with the increasing evidence that higher ventilation rates, when applied appropriately and without recirculation, can reduce SARS-CoV-2 exposure and subsequent infection risk^72–75^. Mechanistically, the overall airborne concentration of virus-containing particles can be reduced through natural or mechanical increases in ventilation rates, thereby reducing the viral dose exposure to a susceptible worker^36^. An important limitation to this intervention has been noted by Lee et al.^73^, who found that even at high ventilation rates of 20 h^-1^, this intervention could not overcome the high infection risk associated with an infected and susceptible worker sustaining contact in small, enclosed office spaces (20 m^3^). This suggests that larger facilities, like the indoor packing facility modeled here (460 m^3^), would likely see a greater reduction in SARS-CoV-2 infection risk upon increasing their ventilation rate, when compared to a smaller facility or office spaces.

Consistent with numerous laboratory^76–78^ and empirical^27,79^ studies testing reduction in risk associated with wearing face masks, our findings for individual indoor and outdoor scenarios, along with the cumulative daily risk, demonstrate that masks are an effective tool for reducing SARS-CoV-2 infection risk. Of the masks described above, double masking (surgical mask followed by cloth mask) provided the greatest reduction in daily risk, followed by one surgical and cloth mask, respectively. Given the increased transmissibility of newer SARS-CoV-2 variants (Delta: B.1.617.2^80^; Omicron: B.1.1.159^81^), the CDC has recently revised masking recommendations and has transitioned from promoting double masking^82^ to now promoting N95 and KN95 respirators^83^, while surgical masks continue to be recommended for industry by OSHA^84^. After incorporating the universal usage of optimally-fit N95 respirators^77^ throughout our model (data not shown), the 24 h daily cumulative risk for an indoor (0.004, 95% UI: 0.001– 0.009) and outdoor (0.018, 95% UI: 0.009–0.039) produce worker was reduced by 99.6% and 96.2%, respectively. Given the documented facemask accessibility issues faced by produce workers at the onset of the pandemic^85,86^, the implementation of N95 respirators, though of superior risk reduction, is likely not feasible in this occupational setting. However, our work supports the promotion of double masking in these occupational scenarios to further minimize infection risk beyond the use of cloth or surgical masks alone.

Our work provides novel insight into the effectiveness of combining vaccination with food industry recommended infection control strategies to mitigate daily SARS-CoV-2 infection risk among essential produce industry workers. As demonstrated with the agent-based modeling studies of Farthing et al.^87^ and Kerr et al.^88^, while individual interventions (e.g., physical distancing, masking, vaccinations, symptomatic testing, etc.) provided modest reductions in the expected number of SARS-CoV-2 infections, implementing multiple interventions simultaneously provided synergistic reductions in expected infections. It is important to note that this multipronged approach assumes absolute compliance with the interventions described; however, the layering of risk reduction practices is not new to the produce industry^89^. As demonstrated by Mogren et al.^90^, the “hurdle-approach” of enforcing multiple risk reduction strategies with variable degrees of compliance can provide greater reductions in risk than a single intervention with poor compliance.

The strengths of our model include: a detailed exposure assessment designed to simulate the daily work scenarios experienced by indoor and outdoor produce workers; model vetting by academic and food industry partners; and a versatile modeling framework that can be leveraged to evaluate scenarios and contexts outside of the food production and processing industry. The design of this model has allowed us to evaluate the impact of global, national, and industry-specific SARS-CoV-2 intervention recommendations within and across multiple scenarios, highlighting their scenario-specific variability and overall effectiveness at reducing infection risk. While these features are not commonly applied to QMRA models and have recently been the focus of expansive agent-based modeling work^88^, this work highlights the capabilities of QMRA modeling to extend beyond one-time interactions between an infectious and susceptible individual.

As for potential limitations, our model relies on the dose-response relationship of SARS-CoV-1 to assess infection risk. Given that the SARS-CoV-2 dose-response parameter(s) have yet to be defined in the literature, this approach has become common practice across multiple SARS-CoV-2 QMRA studies^47,91–93^. A second limitation to the model is our assumption of a uniform distribution for virus concentration across all respiratory particle size classes. While this assumption has been implemented by the aerosol modeling work of Zhang et al.^47^, it is probably not entirely representative of what is happening in real-world situations. This could have led to an artificially increased risk associated with close proximity (≤1 m) exposure to droplets, given their large diameters (100–750 μm) and, by assumption, a higher concentration of infectious SARS-CoV-2 virus. Another limitation is that we assumed discordant vaccination status (e.g., only the susceptible worker had received a vaccination) among the infected and susceptible produce workers modeled. While vaccine hesitancy has been well documented among the food worker population^94,95^, these findings suggest that promoting a single-dose or two-dose vaccine can reduce daily infection risk by roughly 73.0% or 93.0%, respectively. Future work is needed to understand the SARS-CoV-2 vaccination coverage levels within this essential worker community, the findings of which could then be incorporated into the present model to increase the generalizability of our results.

Our model has demonstrated that the elevated daily SARS-CoV-2 infection risk experienced by indoor and outdoor produce workers can be reduced to <1 × 10^−2^ when vaccinations (optimal vaccine efficacy 86–99%) are combined with recommended infection control strategies. Given their consistency in reducing infection risk for both indoor and outdoor produce workers (92.7– 93.0% reduction), vaccinations should continue to be prioritized as an effective means by which to maintain a healthy workforce and reliable global food supply. However, our findings show that vaccinations alone are not sufficient and should be combined with additional infection control interventions (e.g., handwashing, surface disinfection, universal masking, and increased ventilation) implemented by the food industry to further reduce the potential for SARS-CoV-2 transmission. These findings provide additional evidence supporting international (EU OSHA^96^, WHO^20,97,98^), domestic (CDC^99–101^, U.S. OSHA^84,102^), and industry-specific (FDA^103^, AFFI^26^) recommendations for preventing SARS-CoV-2 transmission during food production and processing shifts, and across various day-to-day exposure scenarios (e.g., shared transportation, employer-provided housing, etc.). Taken together, these findings highlight the need for produce industry management to continue promoting vaccination uptake in their workforce combined with supporting the use of food industry recommended infection control strategies when managing SARS CoV-2 transmission in this essential workforce.

## Data Availability

The code developed and utilized throughout this analysis will be available through GitHub upon acceptance. All model parameters in the present work are contained in the manuscript.

## Acknowledgements

The authors would like to thank Dr. Sanjay Gummalla (American Frozen Food Institute), Dr. Lory Reveil (American Frozen Food Institute), and Dr. Max Teplitski (Produce Marketing Association) for their valuable time and input as food and produce production and processing experts and for conducting surveys of facilities.

