## Supporting Information for "Combined Infection Control Interventions Protect the Essential Workforce from Occupationally-Acquired SARS-CoV-2 during Produce Production, Harvesting and Processing Activities"

### Contents

*S1. SARS-CoV-2 QMRA Modeling Parameters* ....................S2
*S2. Scenario-Specific Modeling Parameters & Viral Dose Calculations* ....................S6
*S3. Model Stability Assessment* ....................S11
*S4. Scenario-Specific Variability & Uncertainty Ratios* ....................S12
*S5. Scenario-Specific Spearman Rho Correlation Coefficients* ....................S16
*References* ....................S19

### Content Summary

Pages: 21

Paragraphs: 11

Tables: 5

Figures: 5

Equations: 8

### S1. SARS-CoV-2 QMRA Modeling Parameters

The model parameters used to inform the three SARS-CoV-2 viral transmission pathways (aerosol, droplet, and fomite-mediated) were grouped into three categories and are summarized in **Supporting Table 1A-C**. These classifications include: 1) viral shedding through coughing respiratory events; 2) fomite-mediated transmission and dose-response parameters; and 3) risk mitigation interventions (infection control interventions and vaccination). Additionally, the scenario-specific parameters, such as the volume of space modeled, temperature and relative humidity, baseline air exchange rate (ACH), and fomite-specific viral decay parameters can be found in **Supporting Table 2**.

**Supporting Table 1A.** SARS-CoV-2 Viral Shedding Parameters

| Class | Parameter | Units | Description | Input Values * | Distribution | Reference |
|---|---|---|---|---|---|---|
| **Virus** | | | | | | |
| | $Log_{10}(C_{virus})$ | $^{PFU}/_{mL}$ | Concentration of virus in saliva | 6.80 (6.10, 7.40) | Triangular | [1,2] |
| | $d_{p,c}$ | cm | Particle diameter for coughing event | $6.0x10^{-4}$ ($2.0x10^{-4}$, $4.9x10^{-4}$) | Triangular | [3–5] |
| | $d_{p,b}$ | cm | Particle diameter for breathing event | $8.0x10^{-5}$ ($3.0x10^{-5}$, $2.0x10^{-5}$) | Triangular | [5,6] |
| | $V_{F,b}$ | $^{mL}/_{Breath}$ | Volume fraction associated with droplet diameters 0.6μm-2.2μm | $2.0x10^{-10}$ ($1.1x10^{-10}$, $2.9x10^{-10}$) | Uniform | [5,6] |
| | $V_{F,c}$ | $^{mL}/_{Cough}$ | Volume fraction associated with droplet diameters 2μm–50μm | $2.3x10^{-6}$ ($1.4x10^{-6}$, $2.6x10^{-6}$) | Triangular | [5] |
| | $V_{F,c}$ | $^{mL}/_{Cough}$ | Volume fraction associated with droplet diameters 50μm–60μm | $6.0x10^{-6}$ ($3.5x10^{-6}$, $6.7x10^{-6}$) | Triangular | [5] |
| | $V_{F,c}$ | $^{mL}/_{Cough}$ | Volume fraction associated with droplet diameters 60μm–100μm | $4.9x10^{-6}$ ($1.1x10^{-6}$, $8.4x10^{-6}$) | Triangular | [5] |
| | $V_{F,c}$ | $^{mL}/_{Cough}$ | Volume fraction associated with droplet diameters 100μm–750μm | $6.8x10^{-3}$ ($4.0x10^{-3}$, $7.6x10^{-3}$) | Triangular | [5] |
| | $F_C$ | $^{Cough}/_{hr}$ | Coughing rate per hour | 24.7 (10.0, 39.3) | Uniform | [7] |
| | $F_B$ | $^{Breath}/_{hr}$ | Breathing rate per hour | 1081 (960, 1,200) | Uniform | [8] |

*Values presented as Mean (Min, Max) for Uniform Distributions & Mode (Min, Max) for Triangular Distributions

**Supporting Table 1B.** SARS-CoV-2 Fomite-Mediated Transmission & Risk Assessment Parameters

| Class | Parameter | Units | Description | Input Values * | Distribution | Reference |
|---|---|---|---|---|---|---|
| **Risk** | | | | | | |
| | $Finger_{sa}$ | $m^2$ | Surface area of three fingertips touching fomite surface | $4.2x10^{-4}$ | Point | [9] |
| | $Hand_{sa}$ | $m^2$ | Surface area of two palms | $4.9x10^{-2}$ | Point | [9] |
| | $\lambda_{hand}$ | $min^{-1}$ | Viral decay on hand surface | 1.20 (0.92, 1.47) | Uniform | [10] |
| | Freq.hs | $Contacts/_{min}$ | Frequency of contacts between hands and fomite | 1.0 | Point | Assumed |
| | Freq.hf | $Contacts/_{min}$ | Frequency of contacts between hands and face | 0.8 | Point | [11] |
| | $F_{23}$ | Proportion | Proportion of virus transferred from hand to face | 0.200 (0.063) | Normal | [12] |
| | $L_{dep}$ | Proportion | Deposition fraction of infectious virus into the lungs | 1.00 | Point | Assumed |
| | $I_R$ | $m^3/_{hr}$ | Inhalation rate per hour | 2.40 (1.62, 3.18) | Uniform | [13] |
| | $k_{risk}$ | Unitless | Dose-response parameter | $6.80x10^{-3}$ | Point | [14] |

*Values presented as Mean (Min, Max) for Uniform Distributions and Mean (SD) for Normal Distributions

**Supporting Table 1C.** Risk Mitigation Interventions Assessed for SARS-CoV-2 Transmission

| Class | Parameter | Units | Description | Input Values * | Distribution | Reference |
|---|---|---|---|---|---|---|
| **Interventions** | | | | | | |
| | $HW_{eff}$ | Log reduction | Hand washing efficacy | 2.00 | Point | [15] |
| | $HW_{freq}$ | $HW/hr$ | Frequency of handwashing per hour | 1.00 | Point | Assumed |
| | $SC_{eff}$ | Log reduction | Surface disinfection efficacy | 3.00 | Point | [16] |
| | $SC_{freq}$ | $SC/hr$ | Frequency of surface cleaning per hour | 0.250 | Point | Assumed |
| | $C_{mask(S)}$ | Log reduction | Source cloth mask efficacy | 0.465 (0.310, 0.620) | Uniform | [17–19] |
| | $C_{mask(R)}$ | Percent reduction | Recipient cloth mask efficacy | 0.529 (0.170, 0.887) | Uniform | [17–19] |
| | $S_{mask(S)}$ | Log reduction | Source surgical mask efficacy | 0.478 (0.387, 0.569) | Uniform | [17–19] |
| | $S_{mask(R)}$ | Percent reduction | Recipient surgical mask efficacy | 0.680 (0.370, 0.998) | Uniform | [17–19] |
| | $D_{mask(S)}$ | Log reduction | Source double mask efficacy | 1.08 (0.274, 1.89) | Uniform | [17–19] |
| | $D_{mask(R)}$ | Percent reduction | Recipient double mask efficacy | 0.685 (0.400, 0.968) | Uniform | [17–19] |
| | $V_{Opt}$ | Percent Reduction | Optimal SARS-CoV-2 vaccine efficacy | 0.880 (0.770, 0.990) | Uniform | [20,21] |
| | $V_{Sub}$ | Percent Reduction | Sub-optimal SARS-CoV-2 vaccine efficacy | 0.630 (0.520, 0.740) | Uniform | [22–24] |

*Values presented as Mean (Min, Max) for Uniform Distributions

### S2. Scenario-Specific Modeling Parameters & Viral Dose Calculations

#### S2.1 Shared Car Transportation (Indoor Worker).

SARS-CoV-2 outbreaks during shared transportation have been well documented[25–27]. However, the exact infection risk to a susceptible individual, exposed to an infected individual, in shared transportation, has yet to be quantified. We leveraged the in-vehicle air pollution study of Ott et al.[28] to parameterize this scenario to simulate a 2005 Ford Taurus mid-size car with a cabin volume of 2.6 $m^3$, baseline ACH range of 0.92 $hr^{-1}$ when parked to 4.1 $h^{-1}$ when traveling 50 mph with the windows closed, and a polyester fomite-specific viral decay[29]. The total amount of time spent in this scenario was 2 h to account for transportation to and from the indoor produce processing facility. We assumed that the infected and susceptible worker would not engage in face-to-face contact while in transit such that only the accumulation of aerosolized particles (excluding droplet particles, fomite-mediated transmission) in the vehicle's cabin would be used to estimate the viral infectious dose ($D_{\text{car transportation}}$), as described in the equation below.

$$D_{car\ transportation}(t) = D_{air,car\ transportation}(t)$$

#### S2.2 Indoor Produce Processing Facility & Breakroom (Indoor Worker).

Expanding upon our previous risk assessment model for an indoor processing facility[30], we parameterized this scenario to simulate a facility volume of 460 $m^3$, baseline ACH of 0.1 $h^{-1}$, and a stainless-steel fomite-specific viral decay[29]. We assumed that the infected and susceptible worker would be physically distanced (2 m apart) per industry guidance throughout their work shift[31,32]. The total amount of time spent in this scenario was 11 h, with an additional 1 h spent in an indoor breakroom. The breakroom simulated had a volume of 139 $m^3$ based on the average minimum requirements to accommodate 10 workers in the space at one time, with equivalent

ACH, fomite-specific viral decay, and physical distancing measures as the indoor produce packaging facility. The combined time spent in the indoor produce packaging facility and breakroom was 12 h. The viral exposure dosage for the indoor produce processing facility ($D_{processing}$) and accompanying breakroom ($D_{indoor\ breakroom}$) can be found in the equation below.

$$D_{processing}(t) = D_{air,processing}(t) + D_{hand,processing}(t)$$

$$D_{indoor\ breakroom}(t) = D_{air,indoor\ breakroom}(t) + D_{hand,processing}(t)$$

**S2.3 Private Residential Housing (Indoor Worker).**

We assumed the indoor worker would have private residential housing where they would not share housing with an infected co-worker. As such, this scenario did not contribute any viral dose to a susceptible indoor worker's daily infection risk.

**S2.4 Shared Bus Transportation (Outdoor Worker).**

We leveraged the school bus particulate matter and ventilation study by Chaudhry et al.[33] to parameterize the shared bus scenario to simulate a cabin volume of 81.4 $m^3$, baseline ACH range of 8.3–14.1 $h^{-1}$ when driving under realistic conditions with the windows closed, and a plastic fomite-specific viral decay[29]. The total amount of time spent in this scenario was 2h to account for transportation to and from the outdoor produce harvesting field. We assumed that the infected and susceptible worker would be seated more than 3 m apart while in transit such that only the accumulation of aerosolized particles in the cabin of the bus would be used to estimate the infectious viral dosage ($D_{bus\ transport}$), as described in the equation below.

$$D_{bus\ transport}(t) = D_{air,bus\ transport}(t)$$

**S2.5 Outdoor Produce Harvesting Field & Breakroom (Outdoor Worker).**

The outdoor seafood market SARS-CoV-2 QMRA framework by Zhang et al.[34] was leveraged to parameterize an outdoor produce harvesting field environment, which simulated a volume of 17 $m^3$ surrounding the stainless-steel processing equipment utilized in agricultural fields[35]. Based on average wind speed data documented by Blanco et al.[36] over a 15-year period in the agricultural environment, we assumed a wind speed distribution of 1–3 mph in the outdoor harvesting field. Given that wind speeds are typically measured by anemometers at a 10 m height, we used the log wind profile equation to adjust for average breathing height:

$$w_b = w_{ref} \cdot \frac{\ln\left({}^{h_b}/_{z_0}\right)}{\ln\left({}^{h_{ref}}/_{z_0}\right)}$$

Where $w_b$ is the wind velocity (m/s) calculated at breathing height $h_b$ (2 m), $w_{ref}$ is the known wind velocity (m/s) at the reference height $h_{ref}$ (10 m), and $z_0$ is the roughness length given the agricultural landscape (0.2 m). The resulting wind speeds were then extrapolated to obtain an ACH range of 457–1371 $h^{-1}$ for the outdoor harvesting field. Due to the close contact nature of outdoor agricultural work[37], the infected and susceptible workers were assumed to be facing one another throughout fresh produce harvesting and processing at a 1 m distance. The total amount of time spent working in the agricultural fields was 11 h, with an additional 1 h spent inside of the shared bus "breakroom". Per author (C.M.R.) expert consultation, we utilized the bus transportation scenario to represent the breakroom space for outdoor workers, as this space is commonly used to cool off and eat meals while not working. For this break scenario, however, we assumed that the workers would be situated within 2 m of one another, meaning that they have the potential to be exposed to both aerosols and droplets in this 1 h break scenario. The infectious viral dosage attributed to the outdoor work shift ($D_{field}$) and associated bus "breakroom" ($D_{bus\ breakroom}$) can be found in the equations below.

$$D_{field}(t) = D_{air,field}(t) + D_{hand,field}(t)$$

$$D_{bus\ breakroom}(t) = D_{air,bus\ breakroom}(t) + D_{hand,bus\ breakroom}(t)$$

**S2.6 Shared Residential Housing (Outdoor Worker).**

The shared residential housing scenario was parameterized based on the OSHA[38] minimum guidelines of 50 $ft^2$ per occupant/room for employer-provided housing, the ASHRAE[39] minimum residential ACH of 0.35 $h^{-1}$, and a glass fomite-specific viral decay[29]. We assumed that the infected and susceptible worker would spend 10 h in this scenario, with 2 h spent in close contact (2 m apart) to account for any time spent cooking, cleaning, and sharing the space together. The remining 8 h were designated for sleeping, where the infected worker would only generate virus-containing aerosols through breathing rather than coughing, which could accumulate in the residential space over time. The total amount of time spent in the residential scenario was 10 h.

$$D_{housing}(t) = D_{air,housing}(t) + D_{hand,housing}(t)$$

**Supporting Table 2.** Scenario-Specific Model Parameters

| | Shared Transportation | | Occupational Location | | | Residential Housing |
|---|---|---|---|---|---|---|
| | *Car* | *Bus* | *Indoor Facility* | *Indoor Breakroom* | *Outdoor Field* | *Shared Housing* |
| Total Exposure Time (hr) | 2.00 | 3.00 | 11.0 | 1.00 | 11.0 | 10.0 |
| Room Volume ($m^3$) | 2.60 | 81.4 | 460 | 139 | 17.0 | 34.0 |
| Temperature (°F) | 75.0 | 75.0 | 65.0 | 65.0 | 85.0 | 75.0 |
| Relative Humidity† | Low | Low | Low | Low | High | High |
| Kinematic Viscosity ($m^2/s$) | $1.54 \times 10^{-5}$ | $1.54 \times 10^{-5}$ | $1.49 \times 10^{-5}$ | $1.49 \times 10^{-5}$ | $1.59 \times 10^{-5}$ | $1.54 \times 10^{-5}$ |
| Dynamic Viscosity ($N \cdot s/m^2$) | $1.83 \times 10^{-5}$ | $1.83 \times 10^{-5}$ | $1.80 \times 10^{-5}$ | $1.80 \times 10^{-5}$ | $1.86 \times 10^{-5}$ | $1.83 \times 10^{-5}$ |
| Air Viral Decay (40) ($hr^{-1}$) | 0.806 | 0.806 | 0.180 | 0.180 | 12.5 | 2.78 |
| Baseline ACH‡ (exchange/hr) | 2.55 (0.98-4.10) (28) | 11.1 (8.96-13.4) (33) | 0.10* | 0.10* | 991 (584-1298) (36) | 0.35*(39) |
| Increased ACH‡ (exchange/hr) | 29.1 (6.50-51.7) (28) | 21.6 (10.5-37.3) (33) | 6.0* | 6.0* | N/A | 3.90 (2.08-5.72) (39) |
| Fomite Material | Polyester | Plastic | Stainless steel | Stainless steel | Stainless steel | Glass |
| Fomite Viral Decay (41) ($hr^{-1}$) | 0.222 | 0.143 | 0.178 | 0.178 | 0.178 | 0.179 |
| Fomite Transfer Efficacy (42) | 0.003 (0.001-0.007) | 0.217 (0.001-0.511) | 0.076 (0.017-0.139) | 0.076 (0.017-0.139) | 0.374 (0.056-0.688) | 0.673 (0.176-0.839) |

† High represents a RH of 60% while low represents a RH of 30%
‡ Values represent mean (95% UI)
* Point distribution with no standard deviation
(#) Represents the reference used for the specified parameter / grouping of parameters

### S3. Model Stability Assessment

Model stability was assessed for the baseline (no infection control measures implemented) outdoor harvest field scenario, which was chosen as the representative scenario, by running 10, 100, 1,000, 10,000 100,000, and 1,000,000 Monte Carlo iterations five times, then calculating the average of the median risk estimates[14]. As shown in Supporting Figure 3, there was some spread in the 5 median risk estimates (white diamonds) calculated when compared to the average of these median risk estimates (black line). We determined that there was minimal variability in risk estimates at 10,000 iterations and beyond, and thus 10,000 iterations were used in analyses for all modeled scenarios.

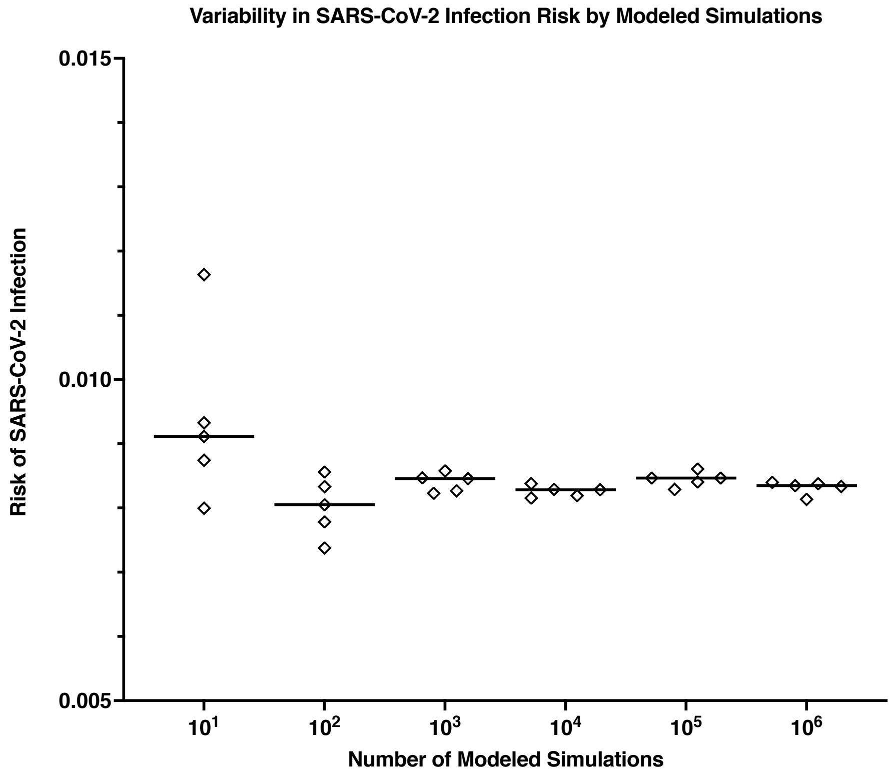


**Supporting Figure 1.** Assessment of model stability in the outdoor harvesting field scenario (12 h risk, 2 m distance) across $10^1$–$10^6$ iterations. Median risk estimates are shown in white diamonds, with the black line representing the average risk estimate across 5 simulations.

**S4. Scenario-Specific Variability & Uncertainty Ratios**

To assess the propagation of variability (natural parameter or distribution heterogeneity) and uncertainty (limited knowledge regarding parameter value(s) or distribution(s)) throughout the modeled scenarios, we leveraged the "mc2d" package in R to perform a two-dimensional Monte Carlo simulation over 10,000 iterations and reported the median and uncertainty interval (UI) comprised of the 2.5 and 97.5 percentiles for the risk estimates. Using the "mcratio" function, the variability, uncertainty, and overall uncertainty (combined impact of parameter variability and uncertainty) ratios were calculated and presented in Supporting Figures 2A-2B and Supporting Table 3. Based on the calculated variability ratios, the parameters which contributed the highest variability to the viral exposure dose were the viral dose transferred from the susceptible worker's hand to their facial mucosa (DT.hh), the Brownian diffusivity calculated in the aerosol-mediated transmission pathway (D), and the viral dose calculated at the designated exposure time. Uncertainty regarding parameter estimates appeared to have a minimal effect on the model outcome (all uncertainty ratios $\leq 1.02$), indicating that variability rather than uncertainty in parameter estimates had a greater impact on the calculated viral exposure dose to a susceptible worker. As future data become available, the parameters utilized in this research can be updated in an effort to reduce the impact of variability and uncertainty on our modeled outcome.

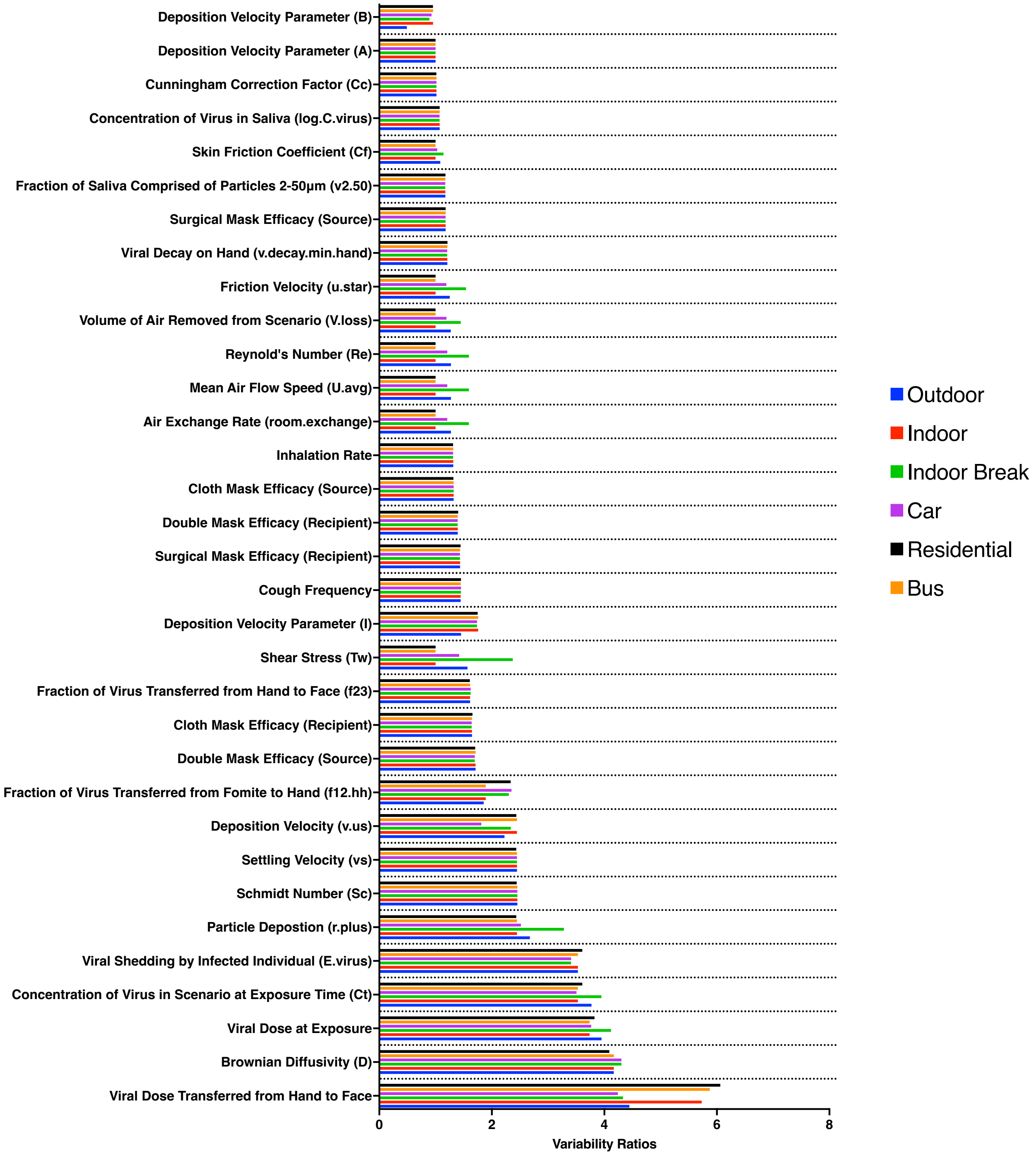


**Supporting Figure 2A.** Variability ratios calculated for the aerosol-, and aerosol-contaminated fomite transmission pathway across each modeled scenario.

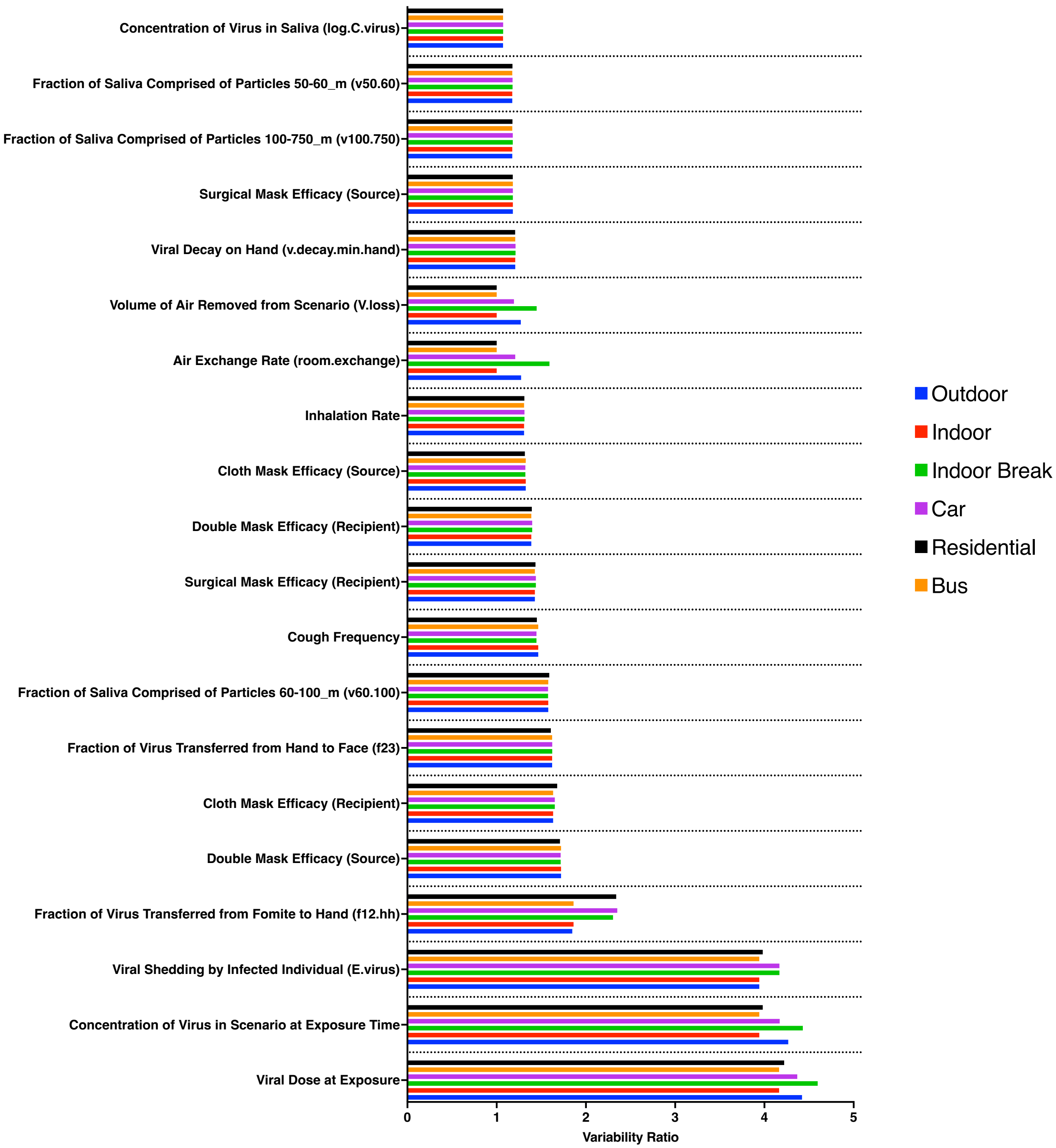


**Supporting Figure 2B.** Variability ratios calculated for the droplet-, and droplet-contaminated fomite transmission pathway across each modeled scenario.

**Supporting Table 3.** Representative Parameter Uncertainty Analysis for the Outdoor Scenario

| Parameter | Variability Ratio[1] | Uncertainty Ratio[1] | Overall Uncertainty Ratio[2] |
|---|---|---|---|
| $Log_{10}(C_{virus})$ | 1.07 | 1.00 | 1.07 |
| v2.50 | 1.17 | 1.00 | 1.18 |
| v50.60 | 1.17 | 1.00 | 1.18 |
| v60.100 | 1.58 | 1.01 | 1.59 |
| v100.750 | 1.18 | 1.00 | 1.18 |
| coughfreq | 1.45 | 1.01 | 1.46 |
| E.virus | 3.50 | 1.02 | 3.58 |
| vs | 2.42 | 1.00 | 2.42 |
| room.exchange | 1.27 | 1.00 | 1.27 |
| U.avg | 1.27 | 1.00 | 1.27 |
| v.us | 2.30 | 1.00 | 2.30 |
| Ct11 | 3.77 | 1.02 | 3.88 |
| f12.hh | 1.84 | 1.01 | 1.86 |
| f23 | 1.62 | 1.01 | 1.63 |
| v.decay.min.hand | 1.21 | 1.00 | 1.21 |
| DT.hh | 4.51 | 1.02 | 4.72 |
| inhalerate | 1.31 | 1.00 | 1.31 |
| V.loss | 1.27 | 1.00 | 1.27 |
| u.star | 1.25 | 1.00 | 1.25 |
| Tw | 1.57 | 1.00 | 1.57 |
| Re | 1.27 | 1.00 | 1.27 |
| B | 0.50 | 1.00 | 0.50 |
| r.plus | 2.65 | 1.00 | 2.65 |
| D | 4.34 | 1.00 | 4.34 |
| Cc | 1.02 | 1.00 | 1.02 |
| Sc | 2.43 | 1.00 | 2.43 |
| A | 1.00 | 1.00 | 1.00 |
| I | 1.45 | 1.00 | 1.45 |
| Cf | 1.08 | 1.00 | 1.08 |
| dosetime11 | 4.00 | 1.02 | 4.12 |

[1] The variability and uncertainty ratios were calculated in the mc2d package in R and represent the impact of a parameter distribution on the model outcome, in this case, the 11h viral exposure dose of SARS-CoV-2 to a susceptible outdoor worker[43].
[2] The overall uncertainty ratio represents the combined effect of uncertainty and variability in a parameter estimate on the model outcome. Lower values represent minimal impact of variability and uncertainty on the final model outcome.

**S5. Scenario-Specific Spearman Rho Correlation Coefficients**

Spearman Rho correlation coefficients were calculated across 10,000 Monte Carlo iterations of each modeled scenario to understand the relationship between each modeled parameter and the viral exposure dose to a susceptible worker. A value greater than 0 indicates that the parameter is positively correlated with the exposure dose, whereas a value less than 0 indicates that the parameter is negatively correlated with the exposure dose. The parameters that were most strongly correlated with the viral dose exposed to a susceptible worker were the total concentration of virus calculated to be in each respective scenario at the designated time (i.e., amount of virus in the indoor processing facility at 11 h), the viral shedding of the infected worker, and concentration of SARS-CoV-2 virus in the saliva of the infected worker. The parameters that were most negatively correlated with the viral dose exposed to the susceptible worker were the double masking efficacy for the infected worker, the surgical mask efficacy of the susceptible worker, and the cloth mask efficacy of the susceptible worker. It is important to note that differences between Spearman Rho values for source vs. recipient protection with the same mask type can be attributed to the differential effectiveness of the masks based on their level of inward and outward protectiveness, described in further detail by Pan et al. 2021[44].

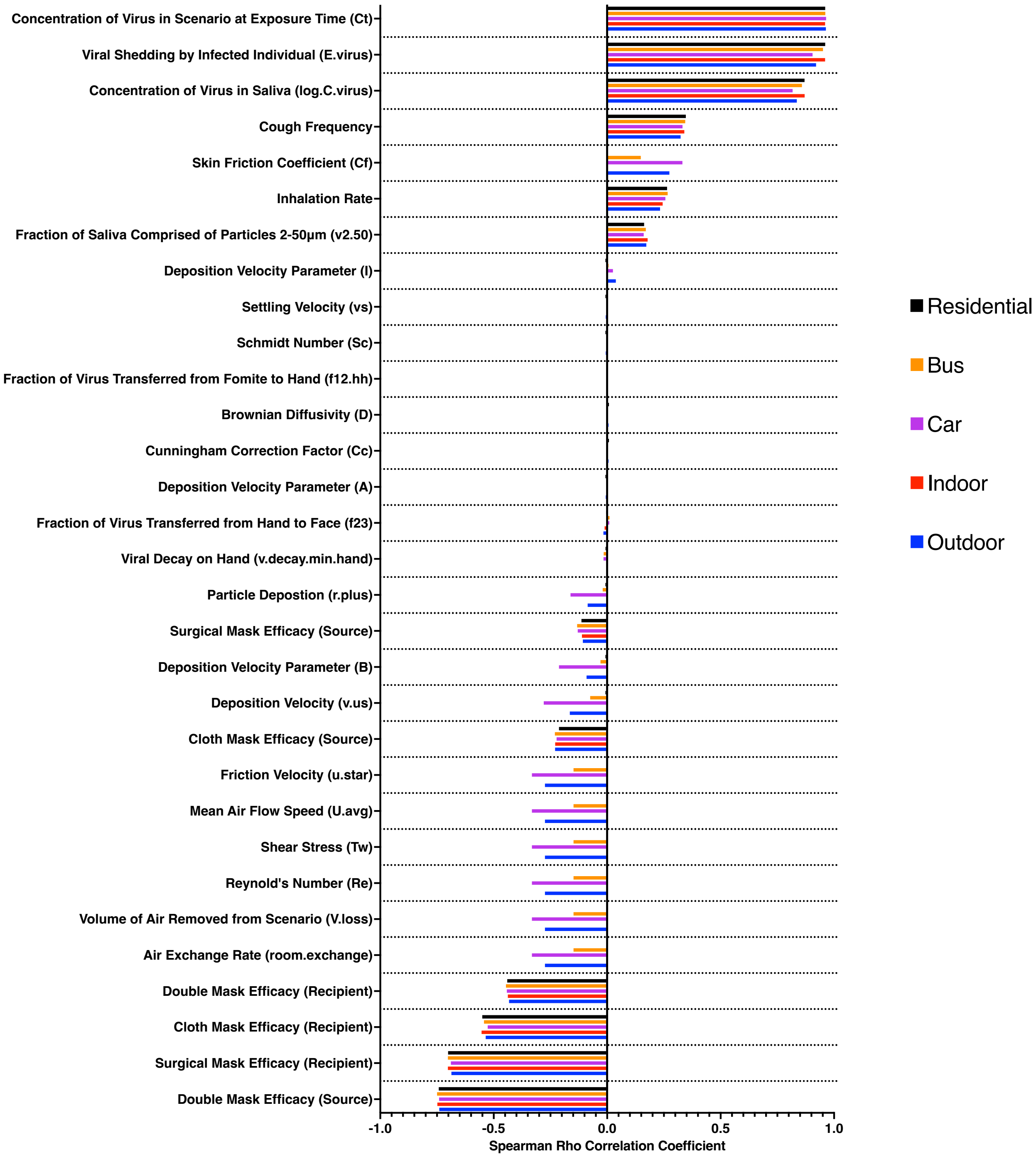


**Supporting Figure 3A.** Spearman rho correlation coefficients calculated for the aerosol-, and aerosol-contaminated fomite transmission viral dose, across each modeled scenario.

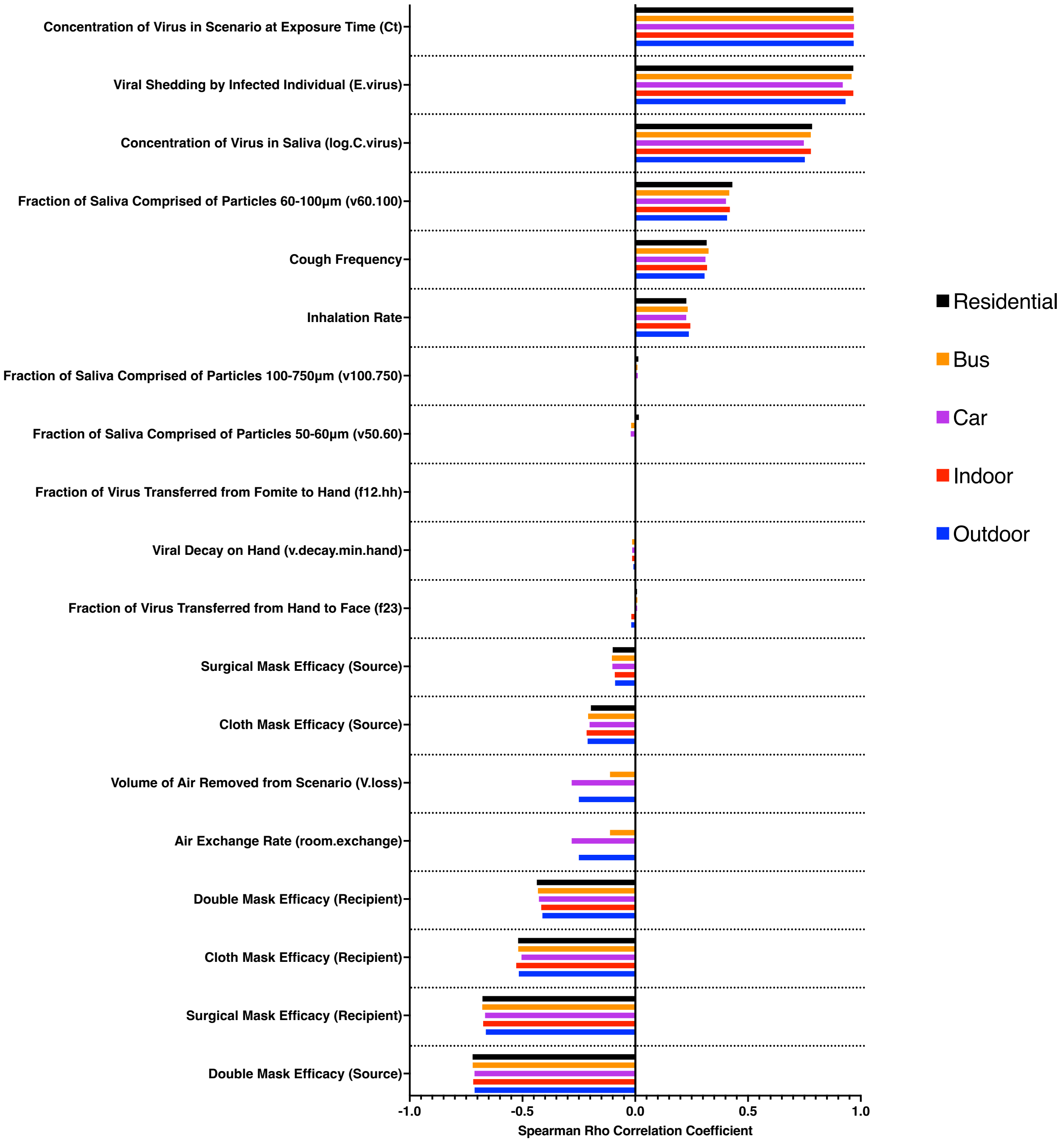


**Supporting Figure 3B.** Spearman rho correlation coefficients calculated for the droplet-, and droplet-contaminated fomite transmission viral dose, across each modeled scenario.
